# Outbreak of a carbapenem-resistant XDR *Acinetobacter baumannii* belonging to the International Clone II (IC2) in a clinical setting in Brazil, 2022

**DOI:** 10.1101/2023.05.12.23289862

**Authors:** Érica Lourenço da Fonseca, Sérgio Mascarenhas Morgado, Fernanda dos Santos Freitas, Priscila P. C. Oliveira, Priscilla M. Monteiro, Lorena S. Lima, Bianca P. Santos, Maria Aparecida R. Sousa, Adriana O. Assunção, Luís Affonso Mascarenhas, Ana Carolina Paulo Vicente

## Abstract

Carbapenem-resistant *Acinetobacter baumannii* (CRAB) is a leading cause of nosocomial infections worldwide, and the occurrence of extensively drug-resistant (XDR) lineages among them is increasing. Most of *A. baumannii* pandemic lineages, known as International clones, are represented by MDR/XDR CRAB strains. The IC2 is considered one of the most successful and widespread pandemic clones, however, it is rare in South America, where IC1, IC4 and IC5 are prevalent. In Brazil, besides sporadic reports, an IC2 outbreak was reported only once in São Paulo city during the COVID-19 pandemics. This study characterized an outbreak caused by IC2 strains (n=16) in a hospital in Rio de Janeiro in 2022. MLST (MLST Pasteur scheme) analysis revealed that all strains recovered from nosocomial infections belonged to ST2 and corresponded to CRAB presenting the XDR phenotype. In general, this broad resistance spectrum was explained by the presence of several antibiotic resistance genes (ARGs) (*armA, bla*_TEM,_ *bla*_OXA-23,_ *bla*_OXA-66_, and *aacA4*-*catB8*-*aadA1*-*qacE*Δ*1/sul1* carried in class 1 integron). Interestingly, the strains characterized here presented a broader resistance spectrum compared to those of the unique other and contemporary IC2 outbreak in Brazil, although they shared most of the ARGs. This study stressed the possibility of the successful establishment of IC2 in Brazilian clinical settings during and after the COVID-19 pandemics in response to a series of events, such as the overuse of antibiotics, during that period.

## INTRODUCTION

*Acinetobacter baumannii* has emerged as one of the most critical opportunistic pathogens for public health worldwide, being frequently involved in ventilator-associated pneumonia, bacteremia, and life-threatening nosocomial infections among severely ill and immunocompromised individuals [1]. This species belongs to the ESKAPE group of pathogens and is featured by its propensity in persisting on inanimate surfaces and acquiring multidrug resistance, which allows it to survive for long periods in the hospital environment. These characteristics contribute to *A. baumannii* person-to-person transmission, and nosocomial spread that favors the successful establishment of infections, leading to a worrisome impact on clinical outcomes [2].

Currently, the golden standard for the treatment of infections caused by *A. baumannii* is carbapenems due to their intrinsic activity against this pathogen [3]. High-risk *A. baumannii* pandemic lineages, named international clones (ICs), generally present a multidrug (MDR) and extensively drug resistance (XDR) character and have been associated with outbreaks around the world. These lineages have mostly been responsible for the spread of carbapenemase genes, such as *bla*_OXA-23_, contributing significantly to carbapenem resistance dispersion [4]. So far, several ICs have been more frequently reported, and among them, the IC2 (represented by the ST2 determined by the MLST Pasteur scheme) is the most globally disseminated and prevalent carbapenem-resistant *A. baumannii* (CRAB) lineage that has been involved with difficult-to-treat outbreaks around the world [5-8]. In Brazil, *A. baumannii* of IC1, IC4, IC5 and to a lesser extent, IC7, have been the most prevalent international clones circulating all over the country [9-14]. Moreover, the IC6 international clone (ST78^PAS^/ST944^OXF^) was recently described in the Brazilian Amazon region [15]. However, in spite of the successful IC2 global spread, its occurrence has barely been reported in Latin America [12,16]. Moreover, IC2 CRAB strains were sporadically described in the Brazilian South region from 1999 to 2002, resurging after ten years (2013-2014). After that, only more recently a local IC2 outbreak was reported in a hospital in São Paulo city [17-21].

Here, we report a recent outbreak caused by CRAB of the IC2 lineage in a hospital in Rio de Janeiro in 2022, and characterize the strains considering their antimicrobial resistance profile, and the presence of genes and elements involved with this phenotype.

## METHODS

From June to September 2022, 16 *A. baumannii* strains were recovered from nosocomial infections occurring in a tertiary care hospital placed in Rio de Janeiro. This hospital has 250 beds of which 60 are divided into six ICUs, all of them with medical, surgical and transplantation units. The bacteria were isolated from several clinical specimens (blood, urine, and bronchoalveolar aspirate) of different inpatients placed in the ICUs. Species identification was obtained by VITEK2 Automated System. The *A. baumannii* identification was confirmed by PCR amplification and Sanger sequencing of *cnp60*, and 16S rRNA genes.

The antimicrobial susceptibility test (AST) was determined by the disc-diffusion method for all antibiotics considered for *Acinetobacter spp*. resistance classification [22], and interpreted according to the Clinical and Laboratory Standards Institute (CLSI) [23]. The Disc-diffusion method was also applied for tigecycline, but interpreted to the breakpoints suggested by the FDA for *Enterobacteriaceae* (susceptible ≥19 mm; intermediate 15–18 mm, and resistant ≤14 mm) (https://www.accessdata.fda.gov/drugsatfda_docs/label/2013/021821s026s031lbl.pdf) [24]. The MIC of polymyxin B and colistin was determined by broth microdilution and interpreted according to the European Committee on Antimicrobial Susceptibility Testing (EUCAST) guidelines (MIC breakpoints for resistance >2mg/L) [25].

The Multilocus Sequence Typing (MLST) based on the Pasteur scheme (PAS) [26] was performed to determine the strains’ sequence typing (ST) and to establish their clonal relationship (https://pubmlst.org/organisms/acinetobacter-baumannii). The presence of the most frequent antimicrobial resistance genes involved with resistance emergence to the clinically relevant antibiotics for *Acinetobacter spp*. [22] (*bla*_IMP_, *bla*_VIM_, *bla*_NDM_, *bla*_OXA-23_, *bla*_OXA-24-like_, *bla*_OXA-58-like_, *bla*_OXA-51-like_, *bla*_GES_, *bla*_CTX-M_, *bla*_TEM_, *armA, rmtD, mcr1-5*) were screened by PCR. The presence of IS*Aba1* upstream *bla*_OXA_ genes was also evaluated. The Quinolone Resistance Determining Region (QRDR) of *gyrA* and *parC* involved with fluoroquinolone resistance emergence in *A. baumannii* was also investigated, as well as the class 1 and 2 integrons and their antibiotic resistance gene content (Table 1).

**Table 1.**
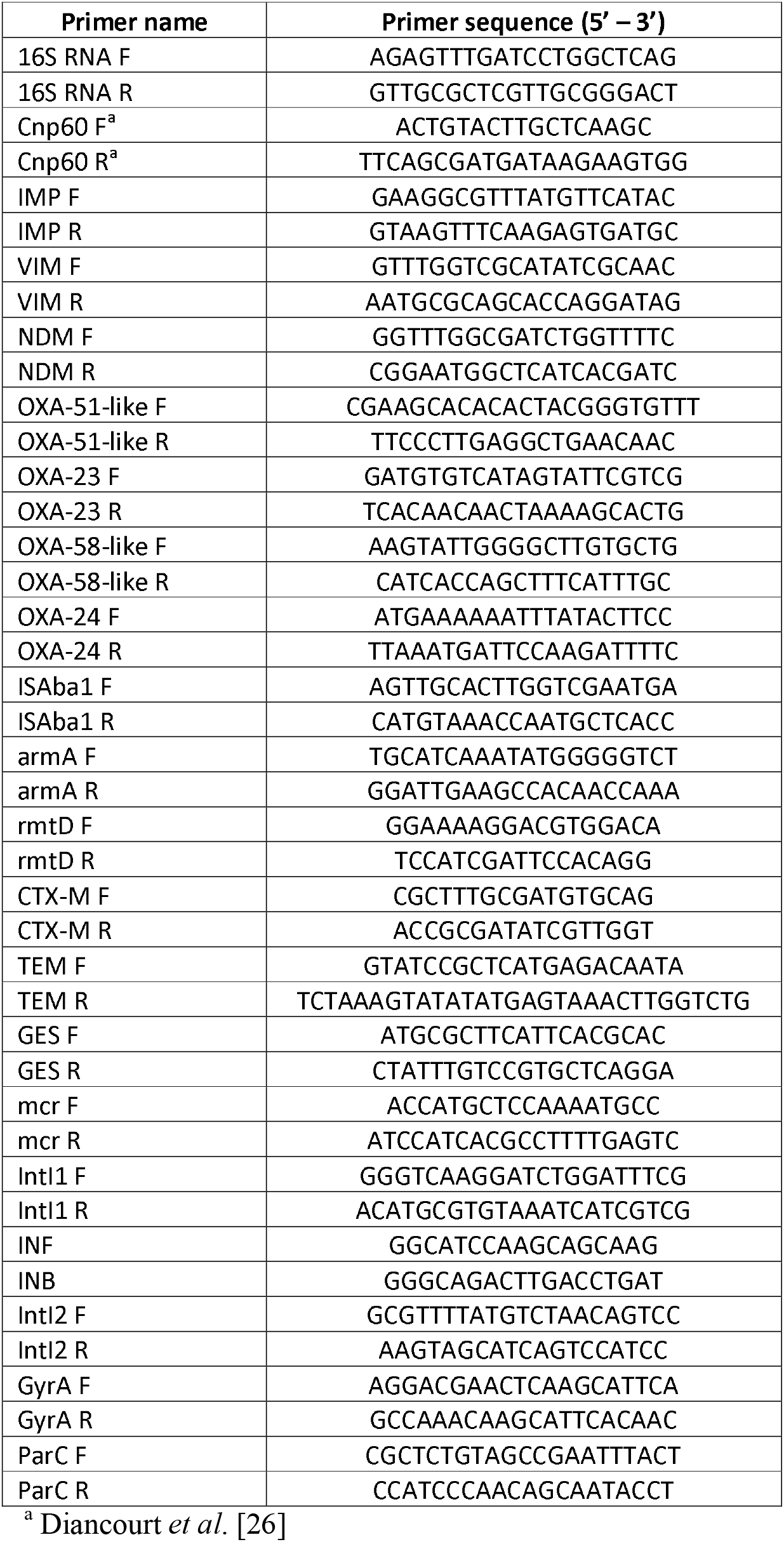
Primers used in this study.

## RESULTS AND DISCUSSION

The MLST analysis revealed that all 16 strains belonged to the international clone IC2. Interestingly, in spite of being a successfully worldwide established clone, the IC2 is sparse and had not been associated with outbreaks in Brazil and South America. After a long period without being detected in Brazil (last reported in 2014) [20], CRAB IC2 resurged in 2020 causing an outbreak in a São Paulo hospital, persisting for at least one year [21]. Interestingly, our study reports the contemporary occurrence of an IC2 outbreak in another Southeast Brazilian state (Rio de Janeiro), however, presenting a broader resistance profile compared with those IC2 strains of the 2020/2021 outbreak [21].

Our results revealed that, in general, all 16 strains presented the extensively drug resistance (XDR) phenotype, with some slight differences in resistance pattern. They were resistant to all cephalosporins and carbapenems, and susceptible to polymyxins (Table 2). Interestingly, the tigecycline resistance profile was heterogeneous among them (resistant=7; susceptible=9; intermediate=3), suggesting differential regulation of genes involved with this resistance. Therefore, it is worth mentioning that despite the XDR phenotype, these strains were treatable since, besides the polymyxins, some strains were also susceptible to minocycline or tigecycline, which increases the chances of favorable outcomes, mainly if these drugs are used in combination [27].

**Table 2.**
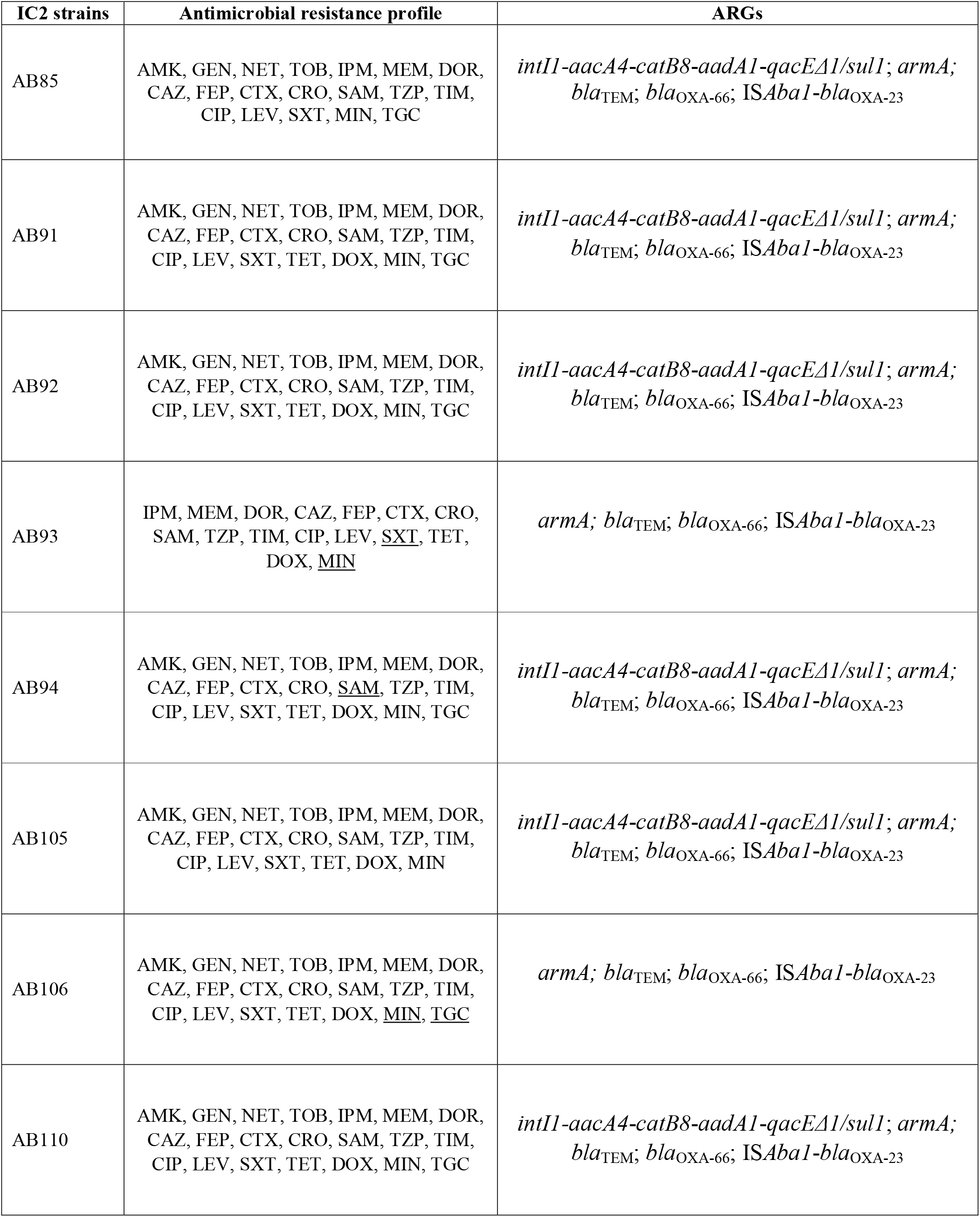
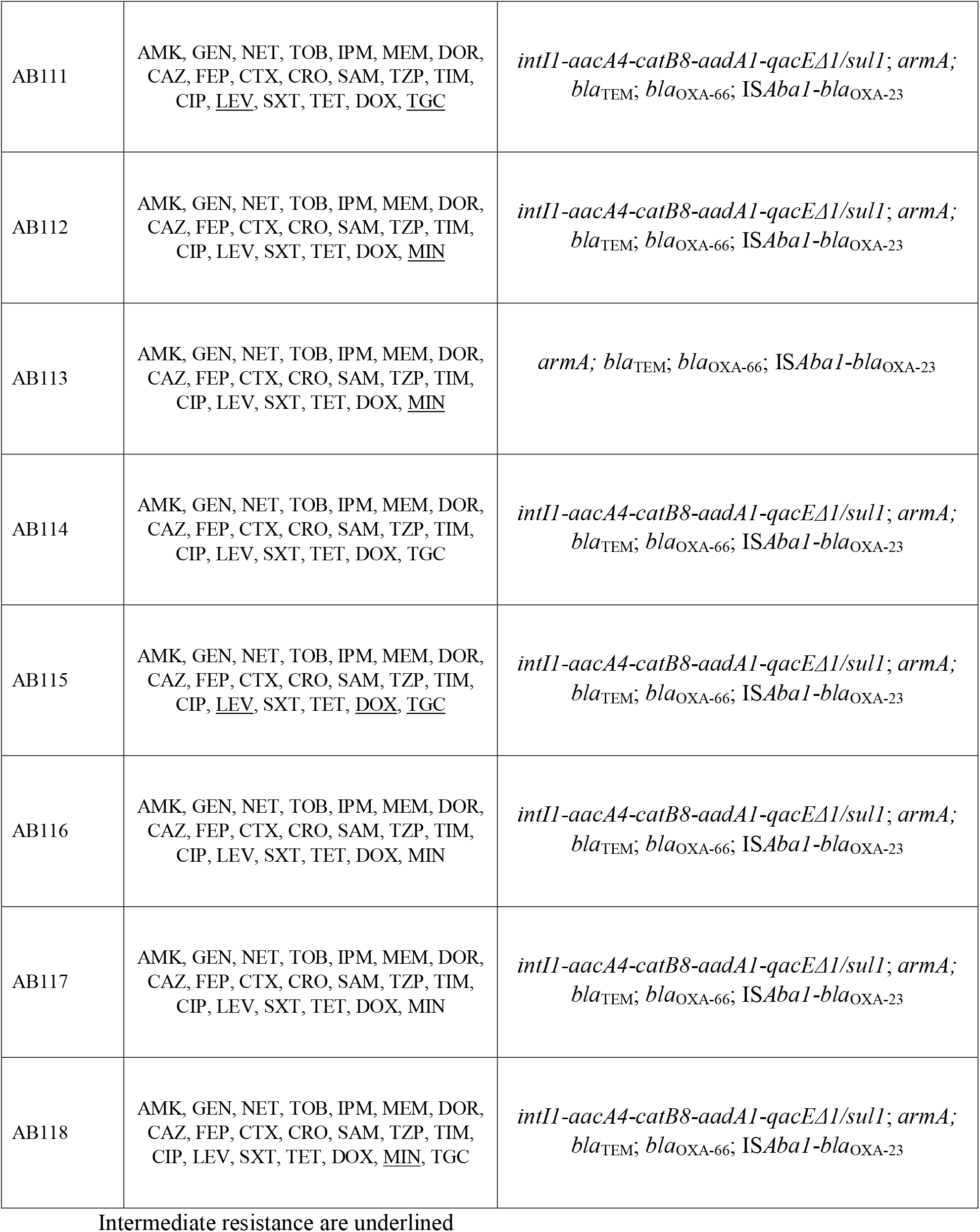
Susceptibility profile and antibiotic resistance genes in the XDR IC2 *A. baumannii* strains of this study

The PCR analyses revealed that most of the strains carried a class 1 integron harbouring the *aacA4*-*catB8*-*aadA1*-*qacE*Δ*1/sul1* gene arrangement, which could contribute to aminoglycoside resistance (*aacA4* and *aadA1*). Besides, the search for ARGs revealed the presence of other genes conferring resistance to aminoglycosides (*armA*), β-lactams (*bla*_TEM_), and carbapenems (*bla*_OXA-23_ and the *bla*_OXA-51_ variant, *bla*_OXA-66_) in all studied ST2 *A. baumannii* (Table 2). All strains harboured the IS*Aba1* upstream *bla*_OXA-23_, which contributed to OXA-23 overexpression. This result demonstrates the important role of *A. baumannii* international clones in OXA-23 and carbapenem resistance dissemination throughout Brazil. Interestingly, IC2 strains incriminated in recent outbreaks among inpatients infected with SARS-CoV-2 in Brazil and Italy [21,28] shared most of the ARGs identified here (*aacA4, aadA1, catB8, armA, bla*_OXA-23_ and *bla*_OXA-66_), indicating that, although transferable, these genes may be well established in the current IC2 lineage circulating worldwide. Moreover, all IC2 strains harboured the Ser83Leu and Ser80Leu substitutions in the GyrA and ParC, respectively, explaining the observed fluoroquinolone resistance.

Camargo *et al*. [21] suggested that the reemergence and occurrence of an IC2 outbreak in São Paulo could be attributed to several unprecedented factors resulting from COVID-19 pandemics, such as the overuse of antibiotics, the high number of inpatients undergoing invasive procedures, such as mechanical ventilation, and drastic alterations in the hospital routine and infrastructure. This was also the case in the studied hospital, and such factors could also have favored the emergence of the IC2 outbreak and the establishment of this international clone in Rio de Janeiro in the COVID-19 post-pandemic period.

Therefore, the present study reported the current occurrence of the CRAB IC2 presenting the XDR phenotype causing an outbreak in a clinical setting in Rio de Janeiro hospital. The expressive ARG set observed in these strains could impact the overall resistance profile of the bacteria circulating in this hospital, due to the possibility of horizontal transfer of these ARGs.

## Data Availability

All data produced in the present work are contained in the manuscript

## AUTHORS’S CONTRIBUTION

Conceptualization, ELF and ACPV; formal analysis, ELF; Collection of data and clinical samples, LAM, PPCO, PMO, LSL, BPS, MARS and AOA; methodology, FSF, NMSB and SMM; writing – original draft, ELF; writing – review and editing, ELF and ACPV; supervision, ACPV.

## ETHICAL STATEMENT

Ethics committee of FIOCRUZ gave ethical approval for this work under the number 39978114.5.0000.5248.

## REFERENCES

1. Appaneal HJ, Lopes VV, LaPlante KL, Caffrey AR. Treatment, Clinical Outcomes, and Predictors of Mortality among a National Cohort of Admitted Patients with Acinetobacter baumannii Infection. Antimicrob Agents Chemother 2022;66:e0197521.

2. Zarrilli R, Pournaras S, Giannouli M, Tsakris A. Global evolution of multidrugresistant Acinetobacter baumannii clonal lineages. Int J Antimicrob Agents 2013;41:11–19.

3. Fragkou PC, Poulakou G, Blizou A, Blizou M, Rapti V et al. The Role of Minocycline in the Treatment of Nosocomial Infections Caused by Multidrug, Extensively Drug and Pandrug Resistant Acinetobacter baumannii: A Systematic Review of Clinical Evidence. Microorganisms 2019;7:159.

4. Karah N, Khalid F, Wai SN, Uhlin BE, Ahmad I. Molecular epidemiology and antimicrobial resistance features of Acinetobacter baumannii clinical isolates from Pakistan. Ann Clin Microbiol Antimicrob 2020;19:2.

5. Trebosc V, Gartenmann S, Tötzl M, Lucchini V, Schellhorn B et al. Dissecting Colistin Resistance Mechanisms in Extensively Drug-Resistant Acinetobacter baumannii Clinical Isolates. mBio 2019;10:e01083–19.

6. Palmieri M, D’Andrea MM, Pelegrin AC, Perrot N, Mirande C et al. Abundance of Colistin-Resistant, OXA-23- and ArmA-Producing Acinetobacter baumannii Belonging to International Clone 2 in Greece. Front Microbiol 2020;11:668.

7. Khuntayaporn P, Kanathum P, Houngsaitong J, Montakantikul P, Thirapanmethee K et al. Predominance of international clone 2 multidrug-resistant Acinetobacter baumannii clinical isolates in Thailand: a nationwide study. Ann Clin Microbiol Antimicrob 2021;20:19.

8. Pascale R, Bussini L, Gaibani P, Bovo F, Fornaro G et al. Carbapenem-resistant bacteria in an intensive care unit during the coronavirus disease 2019 (COVID-19) pandemic: A multicenter before-and-after cross-sectional study. Infect Control Hosp Epidemiol 2022;43:461–466.

9. Caldart RV, Fonseca EL, Freitas F, Rocha L, Vicente AC. Acinetobacter baumannii infections in Amazon Region driven by extensively drug resistant international clones, 2016-2018. Mem Inst Oswaldo Cruz 2019;114:e190232.

10. Fonseca É, Freitas F, Caldart R, Morgado S, Vicente AC. Pyomelanin biosynthetic pathway in pigment-producer strains from the pandemic Acinetobacter baumannii IC-5. Mem Inst Oswaldo Cruz 2020;115:e200371.

11. Camargo CH, Cunha MPV, de Barcellos TAF, Bueno MS, Bertani AMJ et al. Genomic and phenotypic characterisation of antimicrobial resistance in carbapenemresistant Acinetobacter baumannii hyperendemic clones CC1, CC15, CC79 and CC25. Int J Antimicrob Agents 2020;56:106195.

12. Nodari CS, Cayô R, Streling AP, Lei F, Wille J et al. Genomic Analysis of Carbapenem-Resistant Acinetobacter baumannii Isolates Belonging to Major Endemic Clones in South America. Front Microbiol 2020;11:584603.

13. Nodari CS, Fuchs SA, Xanthopoulou K, Cayô R, Seifert H et al. pmrCAB Recombination Events among Colistin-Susceptible and -Resistant Acinetobacter baumannii Clinical Isolates Belonging to International Clone 7. mSphere 2021;6:e0074621.

14. Barcelos Valiatti T, Silva Carvalho T, Fernandes Santos F, Silva Nodari C, Cayô R et al. Spread of multidrug-resistant Acinetobacter baumannii isolates belonging to IC1 and IC5 major clones in Rondônia state. Braz J Microbiol 2022;53:795–799.

15. Fonseca ÉL, Caldart RV, Freitas FS, Morgado SM, Rocha LT et al. Emergence of extensively drug-resistant international clone IC-6 Acinetobacter baumannii carrying blaOXA-72 and blaCTX-M-115 in the Brazilian Amazon region. J Glob Antimicrob Resist 2020;20:18–21.

16. Levy-Blitchtein S, Roca I, Plasencia-Rebata S, Vicente-Taboada W, Velásquez-Pomar J et al. Emergence and spread of carbapenem-resistant Acinetobacter baumannii international clones II and III in Lima, Peru. Emerg Microbes Infect 2018;7:119.

17. Dalla-Costa LM, Coelho JM, Souza HA, Castro ME, Stier CJ et al. Outbreak of carbapenem-resistant Acinetobacter baumannii producing the OXA-23 enzyme in Curitiba, Brazil. J Clin Microbiol 2003;41:3403–3406.

18. Bier KES, Luiz SO, Scheffer MC, Gales AC, Paganini MC et al. Temporal evolution of carbapenem-resistant Acinetobacter baumannii in Curitiba, southern Brazil. Am J Infect Control 2010;38:308–314.

19. Martins N, Dalla-Costa L, Uehara AA, Riley LW, Moreira BM. Emergence of Acinetobacter baumannii international clone II in Brazil: reflection of a global expansion. Infect Genet Evol 2013;20:378–380.

20. Pagano M, Nunes LS, Niada M, Barth AL, Martins AF. Comparative Analysis of Carbapenem-Resistant Acinetobacter baumannii Sequence Types in Southern Brazil: From the First Outbreak (2007-2008) to the Endemic Period (2013-2014). Microb Drug Resist 2019;25:538–542.

21. Camargo CH, Yamada AY, Nagamori FO, de Souza AR, Tiba-Casas MR et al. Clonal spread of ArmA- and OXA-23-coproducing Acinetobacter baumannii International Clone 2 in Brazil during the first wave of the COVID-19 pandemic. J Med Microbiol 2022;71:001509.

22. Magiorakos AP, Srinivasan A, Carey RB, Carmeli Y, Falagas ME et al. Multidrug-resistant, extensively drug-resistant and pandrug-resistant bacteria: an international expert proposal for interim standard definitions for acquired resistance. Clin Microbiol Infect 2012;18:268–281.

23. Clinical and Laboratory Standards Institute. Performance standards for antimicrobial susceptibility testing. Thirty-First Edition. CLSI document M100. Wayne, PA: Clinical and Laboratory Standards Institute; 2021.

24. Food and Drug Administration. FDA Drug Safety Communication: FDA warns of increased risk of death with IV antibacterial Tygacil (tigecycline) and approves new Boxed Warning; 2013. Available from: https://www.fda.gov/drugs/drug-safety-and-availability/fda-drug-safety-communication-fda-warnsincreased-risk-death-iv-antibacterial-tygacil-tigecycline. xAccessed March 3rd, 2023.

25. The European Committee on Antimicrobial Susceptibility Testing. Breakpoint tables for interpretation of MICs and zone diameters (Version 11.0. 2021). https://www.eucast.org/fileadmin/src/media/PDFs/EUCAST_files/Breakpoint_tables/v_11.0_Breakpoint_Tables.pdf [assessed 13 December 2022].

26. Diancourt L, Passet V, Nemec A, Dijkshoorn L, Brisse S. The population structure of Acinetobacter baumannii: expanding multiresistant clones from an ancestral susceptible genetic pool. PLoS One 2010;5:e10034.

27. Fragkou PC, Poulakou G, Blizou A, Blizou M, Rapti V et al. The Role of Minocycline in the Treatment of Nosocomial Infections Caused by Multidrug, Extensively Drug and Pandrug Resistant Acinetobacter baumannii: A Systematic Review of Clinical Evidence. Microorganisms 2019;7:159.

28. Cherubini S, Perilli M, Segatore B, Fazii P, Parruti G et al. Whole-Genome Sequencing of ST2 A. baumannii Causing Bloodstream Infections in COVID-19 Patients. Antibiotics (Basel) 2022;11:955.

